# Analytical performance of 17 commercially available point-of-care tests for CRP to support patient management at lower levels of the health system

**DOI:** 10.1101/2022.04.23.22273766

**Authors:** Serafina Calarco, B. Leticia Fernandez-Carballo, Thomas Keller, Stephan Weber, Meike Jakobi, Patrick Marsall, Nicole Schneiderhan-Marra, Sabine Dittrich

## Abstract

Accurate and precise point-of-care (POC) testing for C-reactive protein (CRP) can help support healthcare providers in the clinical management of patients. Here, we compared the analytical performance of 17 commercially available POC CRP tests to enable more decentralized use of the tool. The following CRP tests were evaluated. Eight quantitative tests: QuikRead go (Aidian), INCLIX (Sugentech), Spinit (Biosurfit), LS4000 (Lansionbio), GS 1200 (Gensure Biotech), Standard F200 (SD Biosensor), Epithod 616 (DxGen), IFP-3000 (Xincheng Biological); and nine semi-quantitative tests: Actim CRP (ACTIM), NADAL Dipstick (nal von minden), NADAL cassette (nal von minden), ALLTEST Dipstick (Hangzhou Alltest Biotech), ALLTEST Cassette cut-off 10-40-80 (Hangzhou Alltest Biotech), ALLTEST Cassette cut-off 10-30 (Hangzhou Alltest Biotech), Biotest (Hangzhou Biotest Biotech), BTNX Quad Line (BTNX), BTNX Tri Line (BTNX). Stored samples (n=660) had previously been tested for CRP using Cobas 8000 Modular analyzer (Roche Diagnostics International AG, Rotkreuz, Switzerland (reference standards). CRP values represented the clinically relevant range (10-100 mg/L) and were grouped into four categories (<10 mg/L, 10–40 mg/L or 10-30 mg/L, 40–80 mg/L or 30-80 mg/L, and > 80mg/L) for majority of the semi-quantitative tests. Among the eight quantitative POC tests evaluated, QuikRead go and Spinit exhibited better agreement with the reference method, showing slopes of 0.963 and 0.921, respectively. Semi-quantitative tests with the four categories showed a poor percentage agreement for the intermediate categories and higher percentage agreement for the lower and upper limit categories. Analytical performance varied considerably for the semi-quantitative tests, especially among the different categories of CRP values. Our findings suggest that quantitative tests might represent the best choice for a variety of use cases, as they can be used across a broad range of CRP categories.

## Introduction

C-reactive protein (CRP) is known to be an acute phase reactant, a glycoprotein produced by the liver and released into the blood stream within a few hours of a tissue injury occurring, at the start of an infection, or due to other sources of inflammation [1]. CRP levels are typically below 3 mg/L in healthy patients, from 10 to 100 mg/L during a mild infection, and as high as 500 mg/L in patients experiencing a severe inflammatory response.

Point-of-care (POC) tests for CRP are increasingly being used in primary care to assist general practitioners (GPs) in the diagnostic workup for various health complaints, including acute cough and abdominal pain, and to differentiate between mild and severe respiratory tract infections [2][3][4]. These tests can be performed using capillary blood samples, with results available within minutes. The use of POC tests for CRP has been widely implemented and is standard practice in many high-income countries to guide the use of treatment for respiratory tract infections, including in Norway and Sweden [5], while in England these tests are recommended by Public Health England and the National Institute for Health and Care Excellence (NICE) [6]. The measurement of CRP values has also proved useful for excluding severe appendicitis or diverticulitis in patients attending with abdominal pain at emergency rooms [7][8][9].

The diagnostic relevance of CRP testing has also been extensively documented for distinguishing between bacterial and viral infections [10][11]. A study form 2013 of Peng, highlighted that CRP levels > 19.6 mg/L (in the serum) might indicated a bacterial infection and guide antibiotic prescription in patient presenting chronic obstructive pulmonary disease [12]. A study form Korppi and coworkers point out that 40 mg/L could be more efficient than 20 mg/L or 80 mg/L for differentiation between viral and bacterial infection, in children with middle or lower respiratory tract infection [13]. These levels provide clinicians with an additional data point, assisting them in forming a correct medical diagnosis. In the Netherlands, for example, a patient’s CRP value must be known before antibiotics can be prescribed to patients with a lower respiratory tract infection [14].

Uncertainty about the cause of an infection can lead to inappropriate antibiotic prescribing, overuse of resources, and disease complications [15] [16]. This is particularly true in low- and middle-income settings, where the presence of wide-ranging etiologies (parasitic, fungal, bacterial, or viral pathogens) and the lack of adequate diagnostic facilities often leads to the choice of therapy being based on empirical knowledge [3][17]. Antibiotics are often dispensed without diagnostic guidance, leading to large quantities of antibiotics being used, which is linked to increasing levels of antimicrobial resistance (AMR) [18][19].

In addition to guiding the use of antibiotics around the world to tackle the growing AMR crisis [17], CRP has recently also been recommended by the World Health Organization (WHO) [20] as a screening tool for tuberculosis (TB) [21]. For this specific use case a cut off of > 5 mg/L was advised because it is the lowest threshold indicating anomaly in many clinical settings [20].

This greater understanding globally of the potential benefits of using CRP in resource-limited settings for different use cases means that POC devices are becoming increasingly important for the global South. As with all diagnostics, the beneficial effects of measuring CRP are linked to the quality of data produced when using the diagnostic product [22][23][24]. Currently, there is limited evidence available as to how well commonly used POC tests perform when compared with central reference laboratory testing [25][26], critical information that is required when planning any expansion of the use of such tests.

The purpose of this study, therefore, is to evaluate the analytical performance of selected POC CRP tests under ideal conditions. We hope that our data will help to guide decision-makers when selecting the most appropriate test for a specific use case. We compared semi-quantitative and quantitative POC tests for CRP with an enzyme-linked immunosorbent assay (ELISA) for CRP and determined the coefficient of variance. As each CRP use-case requires slightly different cut-off values [27], one overarching analysis using a common cut-off value was performed, with the goal of informing programs and interventions across disease areas.

## Materials and methods

### Criteria for selecting tests

We conducted extensive searches of public databases and websites of commercial companies to identify CRP tests currently available on the market. In total, 33 quantitative POC tests, 21 semi-quantitative POC tests, and 2 qualitative lateral flow tests were pre-selected. Predefined go/no go criteria around the availability of tests were then applied to determine whether a particular POC test should be included in the study (Fig 1). The tests that passed this first screening were then evaluated based on cost/market requirements and test characteristics (Fig 1), each criterion was scored and ranked, and tests with the overall highest scores were included in the study. The maximum number of tests to be included in the study was determined by logistical and budgetary considerations. We evaluated eight quantitative tests: QuikRead go (Aidian), INCLIX (Sugentech), Spinit (Biosurfit), LS4000 (Lansionbio), GS 1200 (Gensure Biotech), Standard F200 (SD Biosensor), Epithod 616 (DxGen), and IFP-3000 (Xincheng Biological); and nine semi-quantitative tests: Actim CRP (ACTIM), NADAL Dipstick (Nal von minden), NADAL cassette (Nal von minden), ALLTEST Dipstick (Hangzhou Alltest Biotech), ALLTEST Cassette cut-off 10-40-80 (Hangzhou Alltest Biotech), ALLTEST Cassette cut-off 10-30 (Hangzhou Alltest Biotech), Biotest (Hangzhou Biotest Biotech), BTNX Quad Line (BTNX), and BTNX Tri Line (BTNX). Details about each test are provided in Table 1.

**Fig 1.**
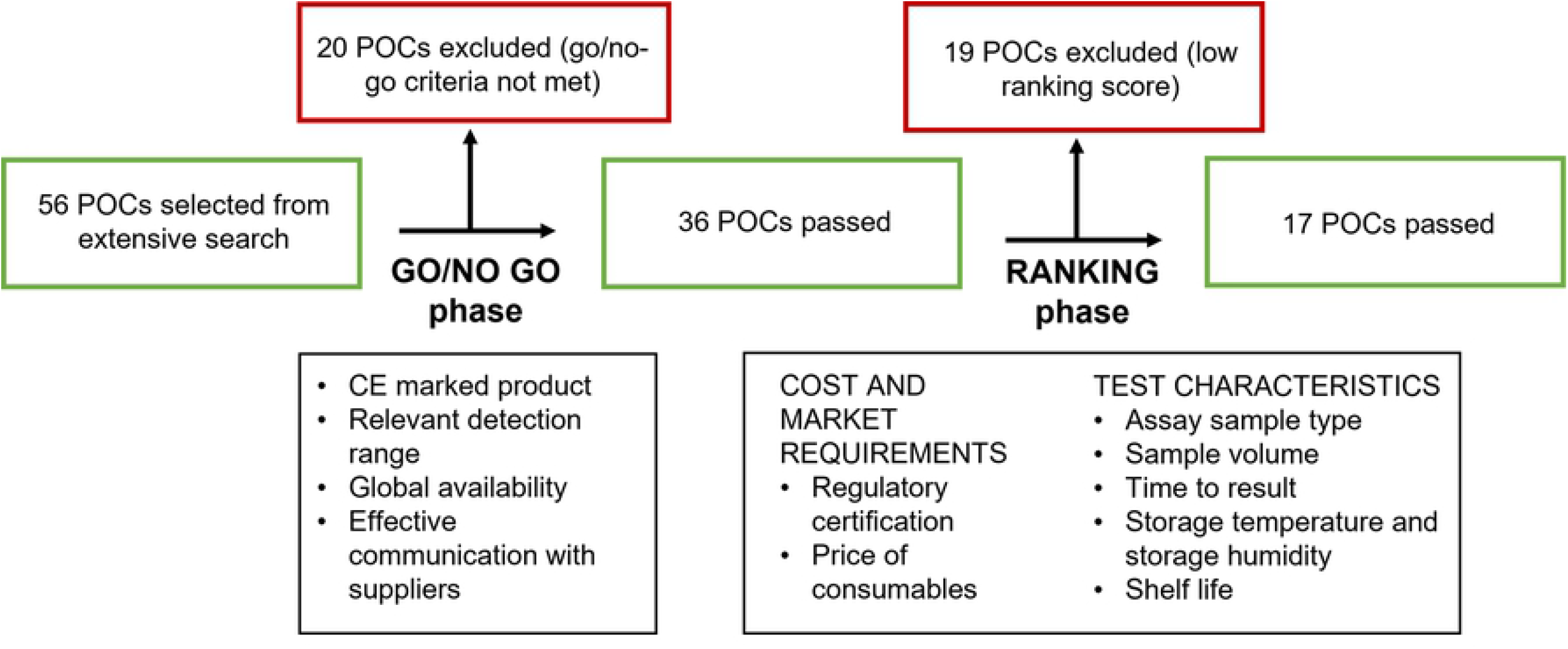
Flowchart for tests selection. Go/no go criteria included both objective and subjective criteria: Whether the test was CE-marked, relevant detection range (from 10 to 100 mg/L CRP for a quantitative test, and one cut-off value of ≥20 mg/L for qualitative and semi-quantitative tests), worldwide distribution, and responsiveness of the manufacturer. Ranking criteria included: Additional regulatory certification (higher score for US Food and Drug Administration-approved products); cost of consumables (lower score for test price >10 USD, higher score for test price <1 USD); assay sample type (lower score for tests requiring serum samples only, higher score for tests that can be used with capillary whole blood, venous blood, or plasma); sample volume (lower score for tests requiring a volume of >50 µL, higher score for tests requiring a volume of <10 µL); time to result (lower score for tests requiring >20 min, higher score for tests requiring <5 min); storage temperature (lower score for tests requiring <4°C, higher score for tests stable up to 40°C); shelf-life (lower score for tests stable for <12 months, higher score for tests stable for >24 months).

**Table 1.** Detailed information on evaluated index tests, A) quantitative and B) semi-quantitative.

| Type of test | Quantitative |  |  |  |  |  |  |  |
| --- | --- | --- | --- | --- | --- | --- | --- | --- |
| **Analyzer name (manufacturer)** | QuikRead go (Aidian) | INCLIX (Sugentech) | SpinIt (Biosurfit) | LS 4000 (LANSIONBIO) | GS1200 (GenSure Biotech) | STANDARD F 200 (SD BIOSENSOR) | EPITHOD 616 (DXGEN) | IFP-3000 (Xincheng Biological) |
| **Analytical range (AR) (mg/L)** | 5–120 (plasma and serum)<br>5–150 (whole blood) | 2.5–300 | 2–300 | 0.5–200 | 0.5–200 | 1–130 (plasma and serum)<br>1–150 (whole blood) | 2.5–200 | 0.5–200 |
| **Sample type** | WB*, serum, plasma | WB, serum, plasma | WB, serum, plasma | WB, serum, plasma | WB, serum, plasma | WB, serum, plasma | WB, serum, plasma | WB, serum, plasma |
| **Sample volume** | 20 µL | 5 µL | 8 µL | 5 µL | 5 µL | 5 µL | 5 µL | 5 µL |
| **Storage temperature for consumables** | 2–8°C | 2–30°C | 2–8°C | 2–30°C | 2–30°C | 2–30°C | 2–10 °C | 2–30°C |
| **Shelf-life of consumables** |  | 24 months | 12 months |  |  | 18 months |  |  |
| **Estimated preanalytical time** | <1 min | 1 min | 5 min | 1 min | 3 min | 3 min | 2 min | 1 min |
| **Time to result** | 2 min | 5 min | 4 min | 2 min | 3 min | 3 min | 10 sec | 3 min |
| **Ability to determine other biomarker(s), yes/no, specification** | Yes<br>CRP+Hb, wrCRP<br>wrCRP+Hb, HbA1c, STREP A, iFOBT, | Yes<br>hsCRP, PCT, Troponin I, HbA1c, β-hCG, Total IgE | Yes<br>BC, HbA1c, COVID-19 Antibody | Yes<br>PCT, SAA, IL-6, HbA1c, PSA, PG I and PG II, TT3, COVID-19 antigen | Yes<br>PCT, SAA, IL-6, HBP, TNF α, Fungal 1,3 β-D glucan, cTn I, NT-proBNP, BNP, cTnI/Myo/CK-MB, HFABP, D-D, MPO, Lp-PLA2, ST2, S100-β, IGFBP-1, PLGF, β-HCG, FSH, LH, PROG, TES, HGH, AMH, INH B, PRL, E2, VB12, FA, 25-OH-VD, SF, TRF, BGP, PTH, PG I, PG II, GP-17, NGAL, Cys C, β2-MG, RBP, MAU, T3, T4, TSH, TG, HbA1c, INS, C-peptide | Yes<br>U-Albumin FIA, HbA1c | Yes<br>U-Albumin, HbA1c, Hb, COVID-19 antigen, COVID-19 IgM/IgG, COVID-19 IgM | Yes<br>HsCRP, PCT, IL-6, SAA, cTnI, MYO/CK-MB/cTnI, NT-proBNP, D-Dimer, COVID-19 IgG/M, COVID-19 Antigen, COVID-19 Neutralizing Antibody |
| **Retail price, consumables (USD)** | <3 | <3 | <6 | <1 | <1 | <3 | <2 | <1 |
| **Retail price, analyzer (USD)** | <1300 | <1000 | <2000 | <600 | <1000 | <2000 | <500 | <500 |
| **Analyzer size (mm)** | 190 x 140 x 80 | 230 x 250 x 250 | n/a | 191 x 84.5 x 45 | n/a | 200 x 240 x 205 | 145 x 210 x 240 | 168 x 70 x 44 |
| **Analyzer weight** | 0.4287 kg | 1.93 kg | n/a | 0.6 kg | n/a | 2.7 kg | 1.8 kg | 0.3 kg |
| **Data export and connectivity** | UBS<br>LIS<sup>b</sup>/HIS<sup>c</sup> | USB (PDF, Excel)<br>Email and LIS | UBS,<br>LIS/HIS | Data transfer via<br>USB not possible | Data transfer as PDF or xls<br>via USB not possible<br>Storage via LIS | UBS,<br>LIS/HIS | USB, Wifi<br>LIS/HIS | Data transfer via USB<br>not possible |
| **Practical aspects of the test <sup>a</sup>** | - Analyzer easy to set up and operate<br>- Many languages available<br>- High-throughput measurements are difficult to perform because of the incubation time of 2 min | -Analyzer easy to handle<br>-“Quick Mode” suitable for testing many samples | - Only operated in standard mode, less suitable for many samples | -Portable device<br>-“Quick Mode” allows processing of many samples.<br>-Low battery capacity<br>-Touchscreen difficult to use with gloves | -Simple interface<br>-Very fast workflow possible due to having six cassette slots | -Only operated in standard mode, less suitable for many samples<br>-Single results cannot be exported | -Digital data export not possible.<br>-Sample preparation with three different reagents (time consuming and comparatively high consumption of pipette tips) | -Fast test processing, suitable for many tests (Quick Mode)<br>-No USB.<br>-Touchscreen keyboard small<br>-Sample preparation complex |

B)
| Type of test | Semi-quantitative |  |  |  |  |  |  |  |  |
| --- | --- | --- | --- | --- | --- | --- | --- | --- | --- |
| Test name (manufacturer) | ACTIM CRP dipstick (ACTIM) | NADAL Dipstick (Nal von minden) | NADAL Cassette (Nal von minden) | ALLTEST Dipstick (Hangzhou Alltest Biotech) | ALLTEST Cassette (Hangzhou Alltest Biotech) | ALLTEST Cassette (Hangzhou Alltest Biotech) | BIOTEST Cassette (Hangzhou Biotest Biotech) | BTNX Quad Line dipstick (BTNX) | BTNX Tri Line Cassette (BTNX) |
| Cut-off (mg/L) | ≥10 – 40 – ≤80 | ≥10 – 40 – ≤80 | ≥10 – 40 – ≤80 | ≥10 – 40 – ≤80 | ≥10 – 40 – ≤80 | ≥10 – ≤30 | ≥10 – 40 – ≤80 | ≥10 – 40 – ≤80 | ≥10 – 30 – ≤80 |
| Sample type | WB, serum, plasma | WB, serum, plasma | WB, serum, plasma | WB, serum, plasma | WB, serum, plasma | WB, serum, plasma | WB, serum, plasma | WB, serum, plasma | WB, serum, plasma |
| Sample volume | 10 µL | 10 µL WB<br>5 µL S/P | 10 µL WB<br>5 µL S/P | 10 µL | 10 µL | 10 µL | 10 µL WB<br>5 µL S/P | 10 µL WB<br>5 µL S/P | 10 µL WB<br>5 µL S/P |
| Storage temperature for consumables | 2–25°C | 2–30°C | 2–30°C | 2–30°C | 2–30°C | 2–30°C | 2–30°C | 2–30°C | 2–30°C |
| Shelf-life of consumables |  |  |  |  |  |  |  |  |  |
| Estimated preanalytical time | 2 min | 2 min | <1 min | 3 min | 5 min | <1 | <1 min | <1 min | 1 min |
| Time to result | 5 min | 5 min | 5 min | 5 min | 5 min | 5 min | 5 min | 5 min | 5 min |
| Retail price, consumables (USD) | <1 | <1 | <1 | <1 | <1 | <1 | <1 | <5 | <5 |
| Practical aspects of the test <sup>a</sup> | -Non-homogeneous test line intensity (sometimes the middle of the line is stronger than the edges and therefore difficult to interpret) | Easily readable band width with homogeneous staining and uniform, consistent color | Low intensity test lines but mostly homogeneous | -First band intensity differs considerably from the control line (assessment difficult)<br>-Band intensity often very non-homogeneous | -Band width non-homogeneous - Variations in color intensity | -Bright red background appears (complicates interpretation of line intensity) | -Good band intensity - Homogeneity partly irregular | -Homogeneous bands<br>-Intense coloration | -Homogeneous bands<br>-Intense band staining |

n/a: not applicable
WB: whole blood
<sup>a</sup> Practical aspects were defined based on subjective observations of test performers using the form shown in Fig S1 to systematically collect
feedback for all tests.
<sup>b</sup> LIS: Laboratory information system141 <sup>c</sup> HIS: Hospital information system

### Sample composition

Serum samples used for this study were selected from the biobank curated by FIND, the global alliance for diagnostics [28] [29]. All patients from whom these samples were obtained gave consent for their samples to be stored in a biobank and used for research into diagnostic tools for fever management. Patients included in this study had fever at presentation and were aged from 2 to 65 years [28]. Standardized guidance for sample transport and storage prior to laboratory evaluation was followed [28], and all samples were preserved under temperature-controlled conditions at -20°C until CRP testing. Reference testing for CRP was conducted as part of the original sample characterization using Cobas 8000 Modular analyzer (Roche Diagnostics International AG, Rotkreuz, Switzerland) [28].

To evaluate the samples’ stability over time and confirm that the previously generated CRP values were reliable, a comparison study was performed (n = 33). Samples were thawed (timepoint 2), and CRP was measured using a highly sensitive ELISA (C-reactive protein high sensitive ELISA, IBL International, Hamburg, Germany, reference method RM2). The resulting measurements were then compared with the original CRP values (timepoint 1) obtained using Cobas 8000 Modular analyzer (Roche Diagnostics International AG, Rotkreuz, Switzerland, reference method RM1). As different methods were used at the two timepoints, the analysis acts as a sample stability and method comparison of RM1 and RM2. A Bland–Altman (BA) plot was used to test for equivalence (two one-sided t-test, TOST), applying predefined +/-8% limits for bias at a +/-90% confidence interval (CI). If necessary, Clinical and Laboratory Standards Institute (CLSI) guidelines EP9 allow separate concentration ranges to be analyzed.

### Sample size

For the quantitative comparison, the number of samples (n = 40) was chosen according to the verification protocol described in CLSI guideline EP09-A3 [30]. For the semi-quantitative tests, a sample size of n = 100 (n = 25 per category, see below) was chosen, to achieve sufficient widths of confidence intervals for the agreement measures used in the study.

### Sample selection and study design

For the evaluation of quantitative tests, samples were selected based on the distribution range of CRP values of 961 samples collected for a previous study [28]. The distribution of the samples had the following deciles (10%.. 90%, in mg/L): 4.4, 10, 20, 30, 45, 65, 85, 113, and 164. As the CLSI guideline requires 40 samples for the quantitative comparison analysis, four samples were randomly selected from each of the following ten ranges (mg/L): >0–4.4, >4.4–10, >10–20, >20–30 >30–45, >45–65, >65–85, >85–113, >113–164, and >164. The samples were organized into four sets of 40 samples per set and the same set was used with two different methods (e.g., set 1 for methods 1 and 2, set 2 for methods 3 and 4, etc.). Samples were measured in duplicate.

For semi-quantitative tests, samples were selected to be equally distributed over all test categories. For example, for a test with four categories (0–10–40–80 mg/L) the following distribution of samples was chosen: 25 samples in the range 0–10 mg/L, 25 samples in the range 10–40 mg/L, 25 samples in the range 40–80 mg/L, and 25 samples in the range >80 mg/L. The samples were organized into five sets of 100 samples per set, and the same set was used for two different methods (e.g., set 1 for methods 1 and 2, set 2 for methods 3 and 4, etc.). If part of the measurement process involved visual inspection by a reader, each sample was measured three times, and the respective cassettes were read by two readers (blinded; six reads per sample overall). At the end, 660 samples were selected for this study out of the 961 available.

### CRP testing procedures

All tests were processed according to the manufacturer’s instructions. Briefly, all components were brought to room temperature, and sample dilution was performed as defined by the manufacturer, using the corresponding buffer provided with the kit. After thorough mixing, the diluted sample was transferred, using the appropriate applicator, to the application field of either a cassette or a test strip. In some cases, the test strip was dipped into the diluted sample (e.g., for a dipstick test). Following the defined incubation time specified by the manufacturer, semi-quantitative tests were independently read and evaluated by two readers, directly after each other, who were blinded to each other’s result. For the assessment of test-line intensity, a grid, ranging from 0 to 10, was used. For the quantitative POC tests, the test device (cassette, strip, etc.) was read in an analyzer supplied with the respective tests, with the CRP concentration directly displayed by the analyzer.

### Statistical analysis

Statistical analyses were performed using SAS software (version 9.4). If the equivalence of the index method with the reference method is considered, i.e., if it is examined whether the bias is near to zero, the estimates (slope of Passing–Bablok (PB) regression or mean bias resulting from a BA plot) are presented with their 90% CI, allowing statements similar to the TOST at an alpha level of 0.05, whereby acceptance criteria of +/-10% were applied, otherwise, 95% CI are presented for estimates.

In terms of kappa values, we present both simple and weighted kappa values. Weighted kappa, on the one hand, accounts for similarity between neighboring categories, thus, allowing for a more comprehensive assessment of agreement in terms of actual concentration values, compared to the binary “black & white” viewpoint of simple kappa evaluation, which on the other hand may be more relevant with respect to the clinical decision making. Moreover, the interpretation of kappa values is often based on the proposals of Landis and Koch [31] and Altman [32], where a kappa of >0.8 is considered very good/almost perfect, >0.6 is good/substantial, and >0.4 is moderate. Applying these criteria, the use of the weighted kappa alone would lead to an overly optimistic interpretation.

### Quantitative tests

#### Method comparison

To compare the index test methods with the reference methods, PB regression [33] and BA plots [34] (relative differences) were applied to all measured values, whereby the means of duplicates measurements have been investigated. Visual inspection was performed to exclude obtrusive outliers [30]. Values outside the limits of the analytical ranges (ARs as reported in table 1) were not included in the calculations but are shown in the respective figures. For PB regression, the slope and intercept were estimated together with the respective 90% CI. A CUSUM (cumulative sum control chart) test was also applied to detect any deviations from linearity.

The BA plots were applied to ranges with a homogeneous distribution of differences. The bias, along with its 90% CI, was estimated using the means of duplicate measurements. The limits of agreement (LoA) were estimated as (95%,95%)-tolerance intervals of the observed differences [35], using the variance estimate obtained from single measurements differences to reflect the precision of a real world measurement in assessment of accuracy as presented by LoA.

#### Precision quantitative tests

For each duplicate, except those out of AR, the precision profile of the percent coefficient of variation (%CV) was examined visually. Repeatability was estimated using a random effects ANOVA, the pooled %CV and its 95% CI were calculated for each test.

### Semi-quantitative tests

#### Method comparison

The reference method measurement results were categorized in the same way as the respective index tests. The percentage agreement of the category within the specific ranges of RM was calculated.

The descriptive statistics (tabulation of raw data, scatterplots, and agreement plots) [36], as well as the analysis (kappa, linear weighted kappa, and percentage agreement for a category related to the respective range of reference values), were performed on the 600 single reads.

#### Reliability of semi-quantitative tests

The agreement of results from the two readers were investigated by simple and linear weighted kappa and percentage concordant and percentage discordant values. In total, 300 paired measurements were evaluated for each index test.

### Quantitative and semi-quantitative tests

#### Binary test results

To compare all tests at a cut-off of 10 mg/L, available for both quantitative and semi-quantitative tests, the binary test results were assessed according to the CLSI guideline EP12 [37], so positive and negative percent agreement (PPA, NPA) were estimated together with their Clopper–Pearson 95% CI.

#### Practical evaluation

In addition to their technical performance, the practical aspects of the various POC CRP tests play a role in the reliability of the CRP result. Therefore, to assess the usability of the different tests, we conducted a practical evaluation in the laboratory. The following aspects were considered: Required sample volume, estimated preanalytical time, and duration of the analysis. Moreover, band width and homogeneity were evaluated for the semi-quantitative tests, as was band intensity, using a reference grade provided by FIND (from 0 to 10). The general usability of quantitative test analyzers was also evaluated, based on the subjective interpretation of the two users. Overall, for each semi-quantitative test, 600 observations were performed, while for each quantitative test, 80 observations were evaluated. The form used to record the results of the practical evaluation is shown in the supporting information (S1 Fig).

## Results

Our search identified 56 tests, which were then assessed according to the predefined criteria (Fig 1). Following this assessment, 17 tests (8 quantitative and 9 semi-quantitative) from 13 companies were included in the study (Table 1).

### Equivalence of reference methods

We were able to confirm equivalence (using TOSTs, with acceptance criteria +/-8%) for the results measured using RM1 (Cobas 8000 Modular analyzer Roche Diagnostic) and RM2 (C-reactive protein high sensitive ELISA, IBL International) for the CRP values in the range >10 mg/L (bias=2.1% (90%CI: -0.0%.. 4.3%, 1 outlier removed). The corresponding BA plot is shown in the supporting information (S2 Fig). Low concentrations of CRP, in the range <10 mg/L, were evaluated separately, according to CLSI EP9 guidelines; for this range, a media bias of -0.48 mg/L (90% CI) must be taken into account, which is acceptable for this study.

### Quantitative tests

#### Method comparison

Both method comparison analysis (PB regression and BA plot) detected considerable (and significant) proportional biases for the majority of the compared tests (PB: slopes from 0.822 to 1.571, BA: bias from -28.0% to 31.7%) (Table 2). For three devices (QuikRead go, Spinit, and INCLIX), an acceptable agreement with the reference method was seen. For the PB regression analysis (Fig 2), the slope was in the 0.91–1.1 range (meaning +/-10%) for QuikRead go and Spinit. In the BA plot analysis (Fig 3), the mean bias was -1.2 (90% CI: -4.7 to 2.3%) and -7.5% (90% CI: 9.3–5.7%) for INCLIX and QuikRead go, respectively. The regression analysis (Fig 2) showed that an overestimation of CRP values occurred with STANDARD F 200, EPITHOD 616, and IFP-3000 when compared with the reference method.

**Table 2.**
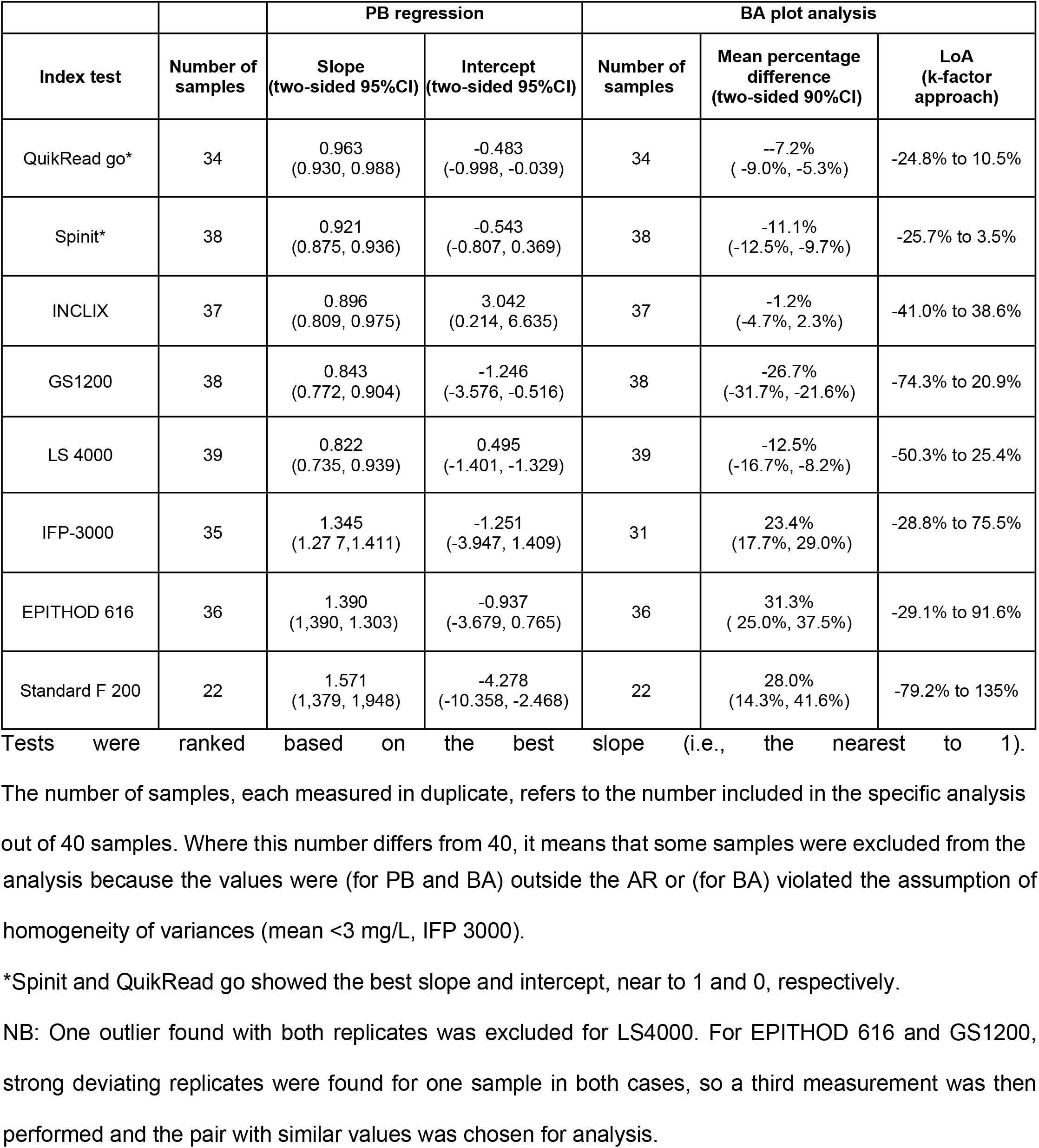
Results for Passing–Bablok regression (slope, intercept) and Bland–Altman plot analysis (percentage bias, limits of agreement (LoA)).

| Index test | Number of samples | PB regression |  | Number of samples | BA plot analysis |  |
| --- | --- | --- | --- | --- | --- | --- |
|  |  | Slope<br>(two-sided 95%CI) | Intercept<br>(two-sided 95%CI) |  | Mean percentage difference<br>(two-sided 90%CI) | LoA<br>(k-factor approach) |
| QuikRead go* | 34 | 0.963<br>(0.930, 0.988) | -0.483<br>(-0.998, -0.039) | 34 | -7.2%<br>(-9.0%, -5.3%) | -24.8% to 10.5% |
| Spinit* | 38 | 0.921<br>(0.875, 0.936) | -0.543<br>(-0.807, 0.369) | 38 | -11.1%<br>(-12.5%, -9.7%) | -25.7% to 3.5% |
| INCLIX | 37 | 0.896<br>(0.809, 0.975) | 3.042<br>(0.214, 6.635) | 37 | -1.2%<br>(-4.7%, 2.3%) | -41.0% to 38.6% |
| GS1200 | 38 | 0.843<br>(0.772, 0.904) | -1.246<br>(-3.576, -0.516) | 38 | -26.7%<br>(-31.7%, -21.6%) | -74.3% to 20.9% |
| LS 4000 | 39 | 0.822<br>(0.735, 0.939) | 0.495<br>(-1.401, -1.329) | 39 | -12.5%<br>(-16.7%, -8.2%) | -50.3% to 25.4% |
| IFP-3000 | 35 | 1.345<br>(1.277, 1.411) | -1.251<br>(-3.947, 1.409) | 31 | 23.4%<br>(17.7%, 29.0%) | -28.8% to 75.5% |
| EPITHOD 616 | 36 | 1.390<br>(1.390, 1.303) | -0.937<br>(-3.679, 0.765) | 36 | 31.3%<br>(25.0%, 37.5%) | -29.1% to 91.6% |
| Standard F 200 | 22 | 1.571<br>(1.379, 1.948) | -4.278<br>(-10.358, -2.468) | 22 | 28.0%<br>(14.3%, 41.6%) | -79.2% to 135% |

Tests were ranked based on the best slope (i.e., the nearest to 1).
The number of samples, each measured in duplicate, refers to the number included in the specific analysis out of 40 samples. Where this number differs from 40, it means that some samples were excluded from the
analysis because the values were (for PB and BA) outside the AR or (for BA) violated the assumption of homogeneity of variances (mean <3 mg/L, IFP 3000).
\*SpinIt and QuikRead go showed the best slope and intercept, near to 1 and 0, respectively.
NB: One outlier found with both replicates was excluded for LS4000. For EPITHOD 616 and GS1200, strong deviating replicates were found for one sample in both cases, so a third measurement was then performed and the pair with similar values was chosen for analysis.

**Fig 2.**
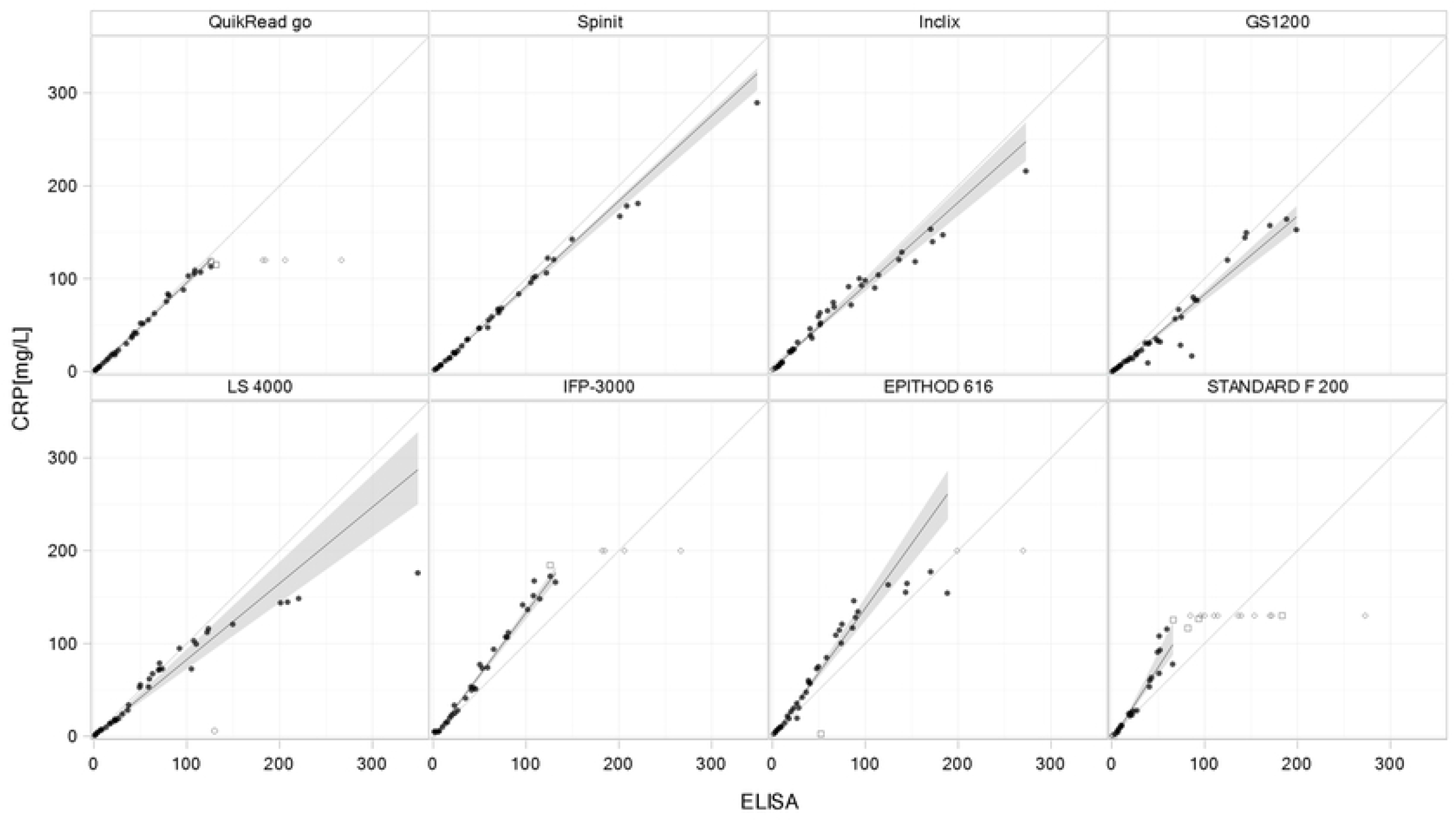
Passing–Bablok regression scatterplots for quantitative tests. Passing–Bablok regression analysis to compare the various quantitative tests and CRP concentrations determined using the RM. Plots are sorted by agreement of the slope with 1, from the upper left to the bottom right. The regression line is represented by a solid black line; dashed gray lines indicate the line of identity. Black dots represent samples included in the analysis, while the other symbols represent samples excluded from the regression analysis measurements (values out of AR and outliers). Gray areas represent the 90% confidence bands, whereby the range covers the observed concentration within AR of the respective index method.

**Fig 3.**
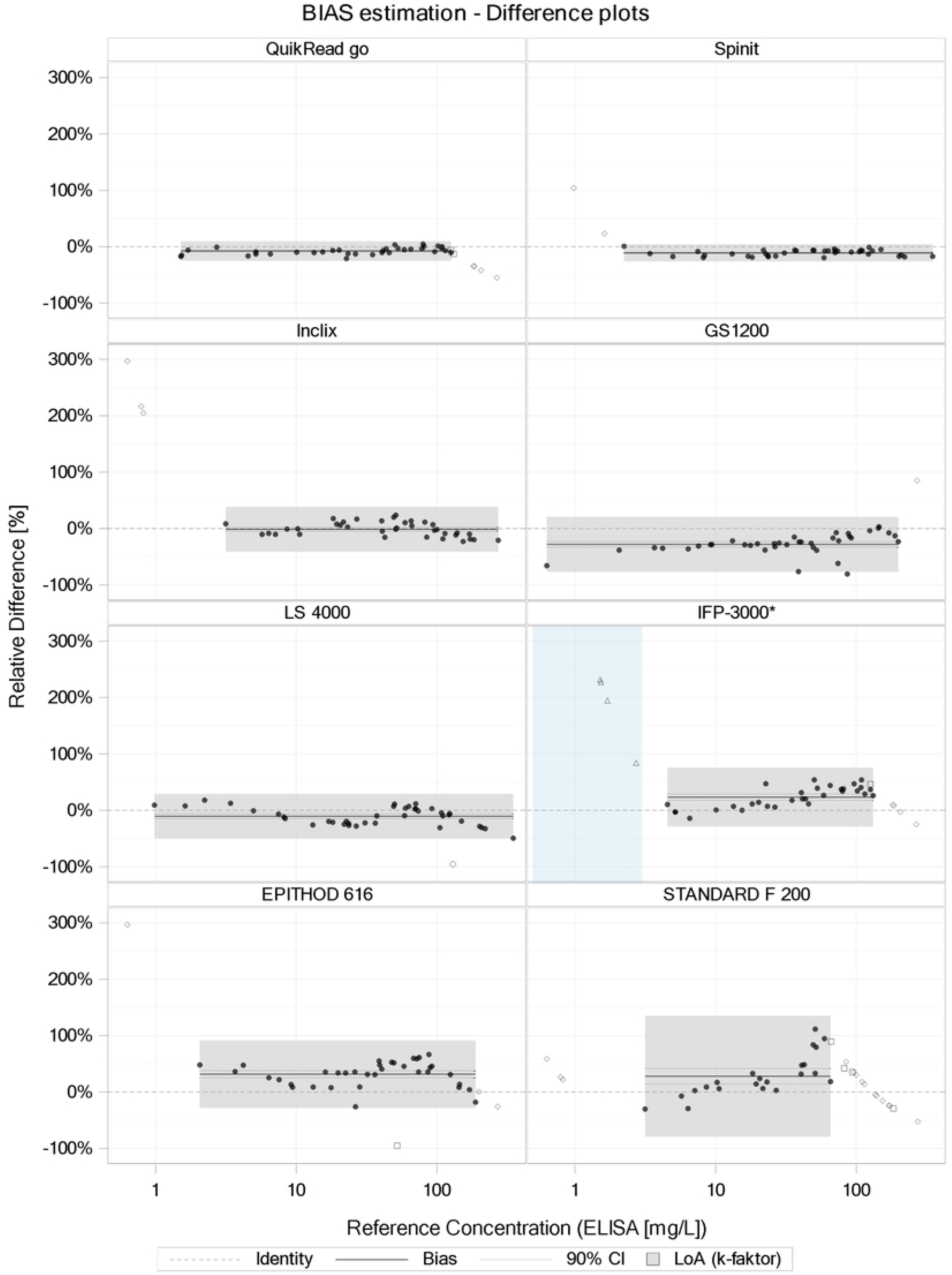
Bland–Altman plots for quantitative tests. Bland–Altman plots comparing CRP POC test results measured using the various quantitative index tests and CRP results measured using the reference method (RM). The X-axes depict the CRP values of the RM, and the Y-axes depict the relative difference between CRP results measured by the POC test under study and the RM. The thick black lines represent the bias and the dotted line its 90% CI; the LoA are represented by the vertical expansion of the gray areas, their horizontal range covers the observed concentrations within AR of the respective index method*. Black dots represent samples included in the analysis, while the other symbols represent excluded data points (values out of AR and outliers). Note: For Standard F200 and IFP-3000, the assumption of concentration-independent relative bias, which is essential for the validity of the BA plot analysis, is questionable. ^*^For IPF3000, three samples were excluded in the low concentration range (<3 mg/L) to achieve constant CV (variance homogeneity) in the analyzed range. As a consequence, the lower limit of the AR is approximately 4 mg/L and not 0.5 mg/L (denoted by the light-blue area in the figure).

Overall, QuikRead go and Spinit (and, to a lesser extent, INCLIX and GS 1200) showed best agreement with the reference method (Fig 2).

#### Precision

The repeatability of the data was analyzed to evaluate the agreement between measured values obtained by replicate measurements (Table 3). The values, expressed as mean % CV, ranged from about 5% up to 16.9%. The tests with the best repeatability were Spinit, LS 4000, and QuikRead go (range approximately 5%), while the largest variability was observed with the SD Biosensor test, with a CV of >15%.

**Table 3.** Repeatability of quantitative tests.

| Repeatability (%CV), 95% CI |  |
| --- | --- |
| Spinit | 4.7% [ 3.8% to 6.0%] |
| LS 4000 | 4.8% [3.9% to 6.1%] |
| QuikRead go | 5.3% [4.3% to 6.9%] |
| IFP-3000 | 9.5% [7.7% to 12.4%] |
| EPITHOD 616 | 13.7% [11.1% to 17.7%] |
| GS1200 | 13.8% [11.3% to 17.8%] |
| INCLIX | 15.4% [12.5% to 19.9%] |
| STANDARD F 200 | 16.9% [13.1% to 23.9%] |

Tests are ranked based on their repeatability, expressed as % CV, 95% CI.

### Semi-quantitative tests

#### Method comparison

The agreement of the semi-quantitative strips with the RM is shown in Table 4. The percentage agreements ranged from high values (100%) to as low as 5.1%, depending on the test and the range. Based on the kappa values, the tests that showed the best agreement were BTNX Quad Line, Biotest, NADAL cassette, and BTNX Tri Line (simple kappa values >0.6). If we also consider the different percentages of agreement for the single categories, the NADAL CRP test and BTNX Quad Line were the two tests that gave the best performance, showing simple kappa values of >0.6 (good agreement) and being the only two tests where we observed a percentage of agreement >50% for all four categories. The Bangdiwala plots (Fig 4), used for analysis visualization, also showed a greater agreement among all the categories for the NADAL cassette and BTNX Quad Line tests.

**Table 4.** Results of the method comparison for semi-quantitative methods.

| Test | Kappa |  | Percentage agreement of the category within the specific range of the reference method |  |  |  |
| --- | --- | --- | --- | --- | --- | --- |
|  | Simple<br>[95%CI] | Weighted<br>[95%CI] | Negative (<10<br>mg/L) | ≥10 to <40 mg/L | ≥40 to <80 mg/L | ≥80 mg/L |
| BTNX Quad Line* | 0.676<br>[0.630, 0.721] | 0.800<br>[0.769, 0.830] | 76.0% (114/150)<br>[68.4%, 82.6%] | 74.7% (112/150)<br>[66.9%, 81.4%] | 56.0% (84/150)<br>[47.7%, 64.1%] | 96.0% (144/150)<br>[91.5%, 98.5%] |
| NADAL cassette* | 0.651<br>[0.604, 0.698] | 0.789<br>[0.759, 0.819] | 88.0% (132/150)<br>[81.7%, 92.7%] | 83.3% (125/150)<br>[76.4%, 88.9%] | 53.3% (80/150)<br>[45.0%, 61.5%] | 70.7% (106/150)<br>[62.7%, 77.8%] |
| Biotest | 0.649<br>[0.603, 0.694] | 0.798<br>[0.770, 0.827] | 99.3% (149/150)<br>[96.3%, 100%] | 45.3% (68/150)<br>[37.2%, 53.7%] | 55.3% (83/150)<br>[47.0%, 63.4%] | 94.7% (142/150)<br>[89.8%, 97.7%] |
| ACTIM | 0.476<br>[0.428, 0.523] | 0.690<br>[0.658, 0.723] | 54.7% (82/150)<br>[46.3%, 62.8%] | 72.0% (108/150)<br>[64.1%, 79.0%] | 16.0% (24/150)<br>[10.5%, 22.9%] | 100% (150/150)<br>[97.6%, 100%] |
| ALLTEST dipstick | 0.443<br>[0.395, 0.491] | 0.671<br>[0.636, 0.707] | 86.7% (130/150)<br>[80.2%, 91.7%] | 37.2% (58/156)<br>[29.6%, 45.3%] | 7.0% (10/142)<br>[ 3.4%, 12.6%] | 100% (150/150)<br>[97.6%, 100%] |
| NADAL dipstick | 0.422<br>[0.373, 0.472] | 0.646<br>[0.610, 0.681] | 98.7% (148/150)<br>[95.3%, 99.8%] | 24.0% (36/150)<br>[17.4%, 31.6%] | 55.3% (83/150)<br>[47.0%, 63.4%] | 48.7% (73/150)<br>[40.4%, 57.0%] |
| ALL TEST cassette | 0.413<br>[0.365, 0.462] | 0.636<br>[0.598, 0.674] | 76.7% (115/150)<br>[69.1%, 83.2%] | 37.2% (58/156)<br>[29.6%, 45.3%] | 11.1% (16/144)<br>[ 6.5%, 17.4%] | 98.0% (147/150)<br>[94.3%, 99.6%] |
|  |  |  | Negative (<10 mg/L) | ≥10 to <30 mg/L | ≥30 to <80 mg/L | ≥80 mg/L |
| BTNX Tri Line | 0.645<br>[0.597, 0.693] | 0.750<br>[0.714, 0.786] | 81.9% (149/182)<br>[75.5%, 87.2%] | 86.6% (123/142)<br>[79.9%, 91.7%] | 80.7% (121/150)<br>[73.4%, 86.7%] | 27.8% (25/90)<br>[18.9%, 38.2%] |
|  |  |  | Negative (<10 mg/L) | ≥10 to <27.7 mg/L | ≥27.7 to <32.5 mg/L | ≥32.5 mg/L |
| ALLTEST cassette | 0.289<br>[0.248, 0.329] | 0.420<br>[0.380, 0.460] | 54.0% (107/198)<br>[46.8%, 61.1%] | 87.8% (158/180)<br>[82.1%, 92.2%] | 11.1% (2/18)<br>[ 1.4%, 34.7%] | 5.1% (10/198)<br>[ 2.4%, 9.1%] |

Kappa (simple, linear weighted) and percentage agreement within specific ranges of the reference method, according to the cut-off of the investigated method. Within equal cut-off groups, tests are ranked based on the simple kappa values.
\*Semi-quantitative tests (BTNX CRP quad line and NADAL cassette) showed a good agreement (simple kappa >0.6) and percentage of agreement for each individual category of >50%.

**Fig 4.**
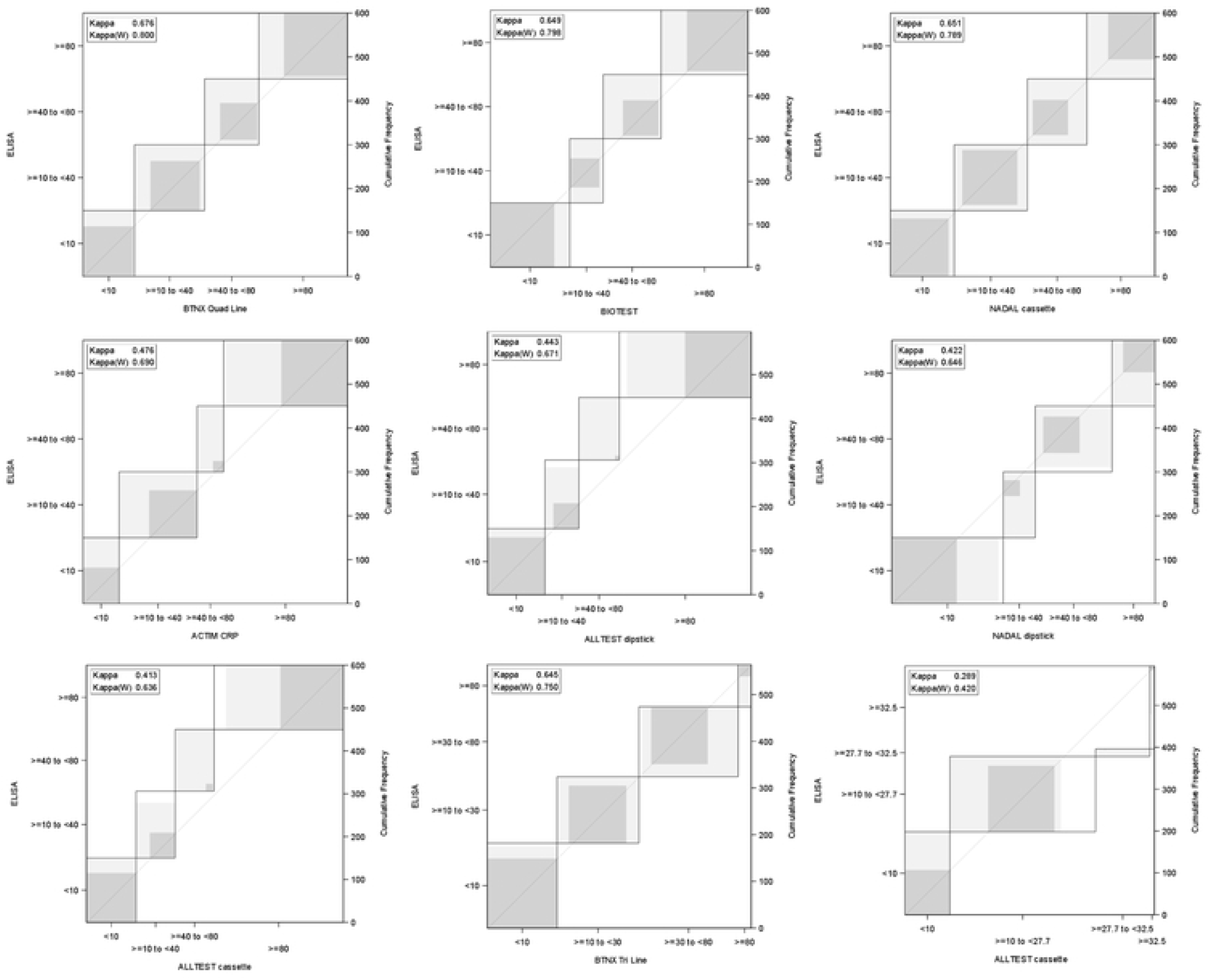
Agreement plots (Bangdiwala plots) for semi-quantitative tests. Y-axis: categories using measurement values of the reference method, X-axis: categories according to the different index tests (units are mg/L). The horizontal width refers to the number of measurement results in the category. The gray shading refers to varying degrees of agreement (darker gray = a better degree of agreement). Overall, a good agreement is indicated by a square shape, with corners on the diagonal identity line and a large amount of dark gray filling the area.

Most of the quantitative tests with four categories showed a lower percentage of agreement for the intermediate categories (10–40 and 40–80 mg/L), while a higher percentage of agreement was observed for the lower and upper categories (0–10 and >80 mg/L) (Table 4). For example, for the cut-off <10 mg/L, we observed among all analyzed tests a percentage of agreement that ranged from 54% to 99%, while for the cut-offs >40 and <80 mg/L, the percentage of agreement was overall quite low, ranging from 11% to 56%. Biotest was the best performing test for the ≤10 mg/L category (99.3% agreement), NADAL cassette for the >10 and <40 mg/L category (83.3% agreement), BTNX Quad Line for the >40 and <80 mg/L category (56% of agreement), and ALLTEST Dipstick for the ≥80 mg/L category, showing 100% agreement.

### Reliability of semi-quantitative tests

The agreement of results provided by the two independent readers is shown in Table 5, overall we could observe a high percentage of concordant results for all tests (from 76.8% to 88.3%). More precisely, BTNX Tri Line, ALLTEST dipstick and BTNX Quad Line tests showed the best proportion of concordant readings, above 85%, and a simple kappa >0.8.

**Table 5.** Between-reader agreement for the semi-quantitative tests.

| Index test | Kappa (simple)<br>[95%CI] | Kappa (weighted*)<br>[95%CI] | Concordant (n/N)<br>[95%CI] | Discordant (n/N)<br>[95%CI] |
| --- | --- | --- | --- | --- |
| BTNX Tri Line | 0.831<br>[0.778, 0.884] | 0.880<br>[0.843, 0.917] | 88.3% (249/282)<br>[84.0%, 91.8%] | 11.7% (33/282)<br>[ 8.2%, 16.0%] |
| ALLTEST Dipstick | 0.818<br>[0.764, 0.873] | 0.907<br>[0.877, 0.937] | 88.0% (263/299)<br>[83.7%, 91.4%] | 12.0% (36/299)<br>[ 8.6%, 16.3%] |
| BTNX Quad Line | 0.811<br>[0.758, 0.863] | 0.889<br>[0.856, 0.921] | 86.0% (258/300)<br>[81.6%, 89.7%] | 14.0% (42/300)<br>[10.3%, 18.4%] |
| BIOTEST | 0.752<br>[0.693, 0.810] | 0.868<br>[0.835, 0.901] | 82.0% (246/300)<br>[77.2%, 86.2%] | 18.0% (54/300)<br>[13.8%, 22.8%] |
| ALLTEST Cassette<br>(four categories) | 0.734<br>[0.673, 0.796] | 0.861<br>[0.825, 0.896] | 82.3% (247/300)<br>[77.5%, 86.5%] | 17.7% (53/300)<br>[13.5%, 22. 5%] |
| NADAL Cassette | 0.729<br>[0.669, 0.789] | 0.835<br>[0.796, 0.873] | 79.7% (239/300)<br>[74.7%, 84.1%] | 20.3% (61/300)<br>[15.9%, 25.3%] |
| ACTIM CRP | 0.725<br>[0.663, 0.787] | 0.848<br>[0.811, 0.885] | 81.7% (245/300)<br>[76.8%, 85.9%] | 18.3% (55/300)<br>[14.1%, 23.2%] |
| NADAL Dipstick | 0.674<br>[0.609, 0.739] | 0.817<br>[0.779, 0.856] | 77.3% (232/300)<br>[72.2%, 81.9%] | 22.7% (68/300)<br>[18.1%, 27.8%] |
| ALLTEST Cassette<br>(three categories) | 0.599<br>[0.516, 0.682] | 0.650<br>[0.575, 0.724] | 76.8% (228/297)<br>[71.5%, 81.5%] | 23.2% (69/297)<br>[18.5%, 28.5%] |

Simple kappa, linear weighted kappa, and the percentages of concordant and discordant results are shown, including the 95% CI. The tests are ranked according to the simple kappa results. For each test, 300 measurements (100 samples in triplicate) were evaluated, except for the ALLTEST dipstick (1 of 300 was not evaluable) and the ALLTEST cassette (3 of 300 were not evaluable).

### Quantitative and semi-quantitative tests

#### Method comparison of binary test results

To allow comparison across all the tests and hence aid the selection for different use cases, one cut-off value was chosen (10 mg/L) that could be found across all of the selected tests (quantitative and semi-quantitative) (Fig 5). Compared with semi-quantitative methods, the PPA (positive percent of agreement) and NPA (negative percent of agreement) for the quantitative tests were more accurate at the selected cut-off; in fact, all PPAs were more than 90% and all NPAs were 100%, with the exception of the EPITHOD 616 test (NPA = 75%). For the semi-quantitative tests, the PPAs were similarly high to those seen with the quantitative tests (except 76.4% for the NADAL Dipstick test), but lower NPAs were observed (two tests, 50%–60%; two tests, 60%–80%; and three tests, 80%–90%), while only two tests showed an NPA >90% (NADAL Dipstick and BIOTEST). This means that for a use case requiring a cut-off of 10 mg/L, a quantitative test is recommended.

**Fig 5.**
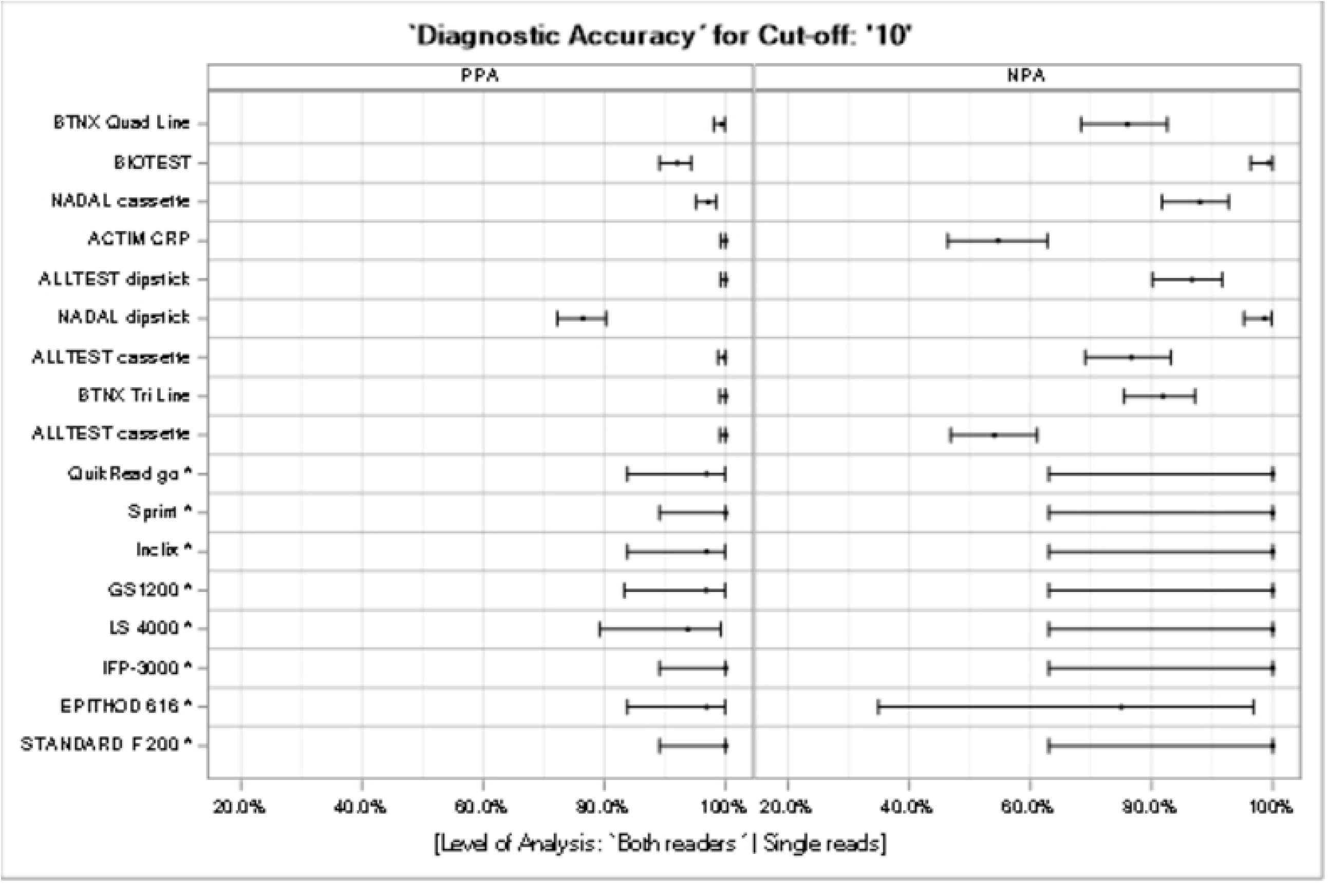
PPA and NPA for semi-quantitative and quantitative tests (cut-off: 10 mg/L). For a cut-off of 10 mg/L, which was available for all quantitative and semi-quantitative methods, the binary test results were assessed; thus, the positive and negative percent agreement were estimated, together with their 95%-Pearson Clopper CI.

## Discussion

Here, we report the results of a method comparison analysis, in which eight quantitative and nine semi-quantitative tests used to measure CRP levels were evaluated for their analytical performance, by comparing them with a known reference method following standard guidelines [38]. To the best of our knowledge, this is the first study where this number of POC tests for CRP have been compared and the results made publicly available to assist developers, users, and procurers. The experiments were designed to cover the expected clinical range of CRP values, with a particular view toward the use case of guiding antibiotic prescribing and other triage decisions at the POC [28] [20] for a broad range of CRP values, in line with the distribution spectrum of CRP values measured in clinical samples. Overall, the quantitative tests showed a satisfactory performance, with the QuikRead go and Spinit tests (and, to a lesser extent, the INCLIX and GS 1200 tests) displaying better agreement with the reference method than the other quantitative tests. For most of the semi-quantitative tests, the percentage of agreement with the reference method varied according to the CRP-level category being examined. Notably, tests with three test bands (the equivalent of four categories) showed a lower percentage of agreement for the 10–40 and 40–80 mg/L categories (from 7% to 83%), while a higher percentage of agreement was observed for the CRP categories <10 (from 54% to 99%) and >80 mg/L (from 48% to 100%). For BTNX Tri Line, on the contrary, a high percentage agreement was observed for the intermediate categories 10–30 and 30–80 mg/L (86 % and 80% respectively), while the category > 80 mg/L showed only 27% of agreement. The current results highlight that, depending on the relevant CRP cut-off needed for the clinical use-case, the utility of the different tests may vary.

The (binary) diagnostic accuracy for CRP-measurements at a cut-off 10 mg/L was also explored, as this is a common cut-off used, across tests and use cases, like tuberculosis and pneumonia [39][21]. While differences existed, overall, the findings suggested that all of the quantitative tests could be used for the cut-off value of 10 mg/L and, more in general, for a broad clinical relevant range, while none of the semi-quantitative tests performed well, and a more cut-off/range-specific selection will be needed. The latter is a critical finding, considering the current push for decentralized testing to inform antibiotic use, TB triage, or other clinical interventions at the first point of contact in resource-limited settings [21][40]. While simple lateral flow-type semi-quantitative tests are likely considered easier to perform, cheaper and closer to existing target product profiles [16], our finding highlights that quantitative tests might have a broader utility across a wider range of CRP categories and hence use cases.

Our findings align with those of a previous study conducted by Minnaard and colleagues, in which the analytical performance of QuikRead go was evaluated [25]. Although we cannot directly compare our results, as in their BA analysis they calculated the mean differences for three ranges of CRP (<20, 20–100, and >100 mg/L), their study showed a good agreement with the reference method for QuikRead go. In contrast, an evaluation conducted by Brouwer and colleagues [26], also using QuikRead go, revealed a significant underestimation of the CRP value compared with the reference method (slope = 0.85), while for our study the lower values provided by QuikRead go were deemed acceptable (slope =0.96). In accordance with our results, their evaluation of the performance of the semi-quantitative test ACTIM also showed an insufficient correlation with the reference method. Overestimation as well as underestimation of CRP values has been observed [26].

Although we aimed to follow the standardized procedures as outlined by the CLSI [37][30], there were some limitations to our study. Two different reference methods were used (C-reactive protein high sensitive ELISA from IBL International and Cobas 8000 Modular analyzer from Roche Diagnostics International AG), but we could confirm equivalence within the prespecified limits, hence it should not be assumed that this caused any issues. Regarding the diagnostic accuracy at 10 mg/L, few samples were used in the range <10 mg/L for the quantitative tests. This is because the focus of this study was a method comparison analysis, and samples were selected to cover a broad range of CRP values.

In summary, we set out to provide pragmatic data to users, procurement agencies, laboratories, implementers, and ministries of health, which can help to inform access to and roll-out of CRP testing tools for a variety of use cases, outside of central reference laboratories. CRP appears on the Essential Diagnostics List produced by WHO and those of individual countries [41][42] and is recognized to be a simple (albeit imperfect) tool to complement clinical assessments to guide antibiotic treatment [43] or conduct triage for infectious diseases [21]. Therefore, it is now of critical importance to continuously assess relevant tools and identify diagnostic tests that can ensure quality data are being generated and used for patient care at all levels of the health system.

## Supporting information

Fifure S2 (supporting info)

## Data Availability

All relevant data are within the manuscript and its Supporting Information files

## Acknowledgments

The authors wish to thank the participants who provided the samples and Adam Bodley for medical writing assistance and the editorial support.

## Supporting information captions

**S1 Fig. Practical evaluation template**

**S2 Fig. Bland–Altmann plot for the stability of reference measurements**. A) excluding values <10 mg/L;

B) values from 0 to 10 mg/L. Gray solid line: bias, grey dotted line: 90% confidence interval band. Gray dash line: allowable range. Symbol star: outlier not included in analysis.

