## Supplementary figures and images for "Analytical performance of 17 commercially available point-of-care tests for CRP to support patient management at lower levels of the health system"

### Fifure S2 (supporting info)

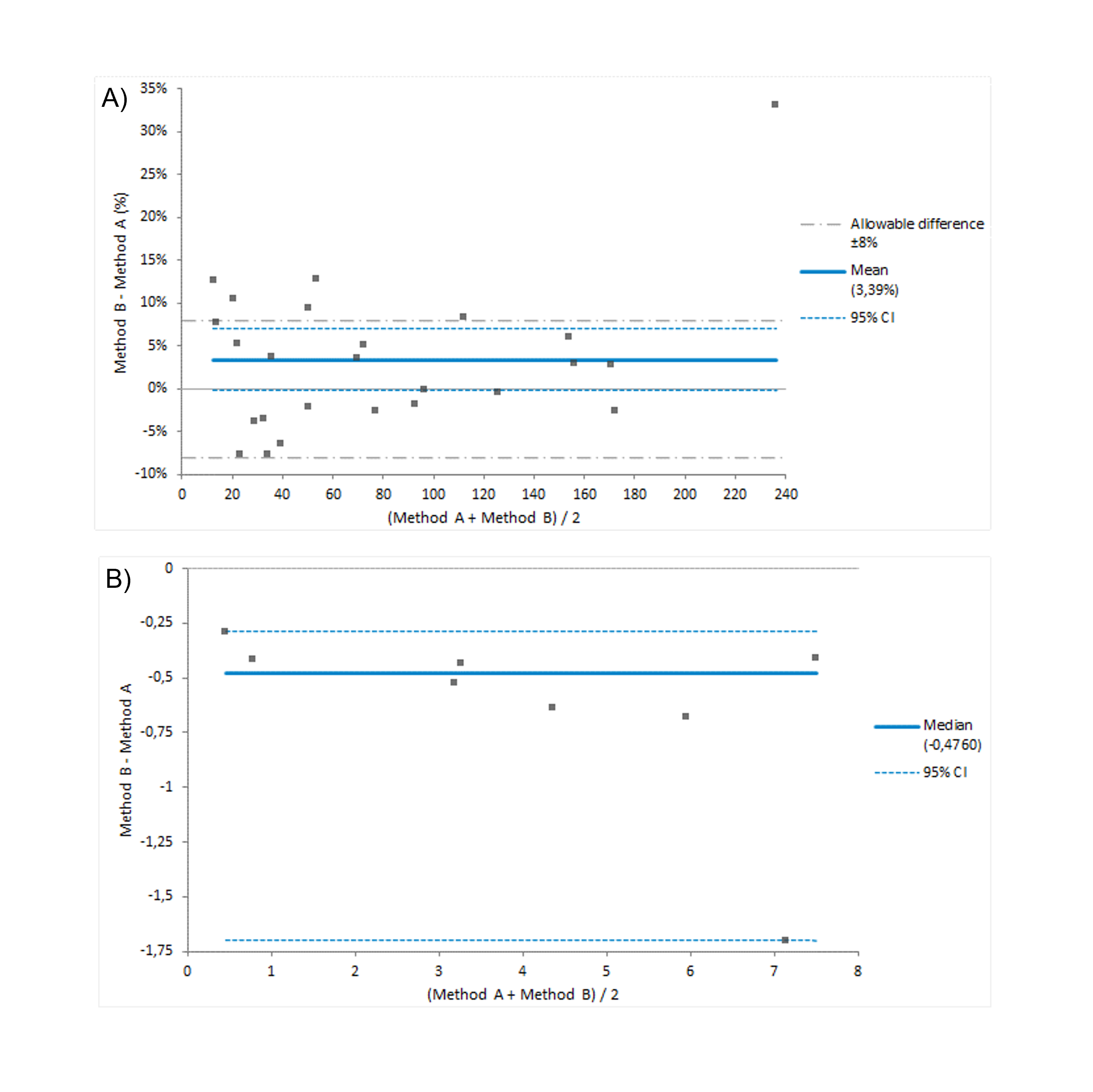

### Figure S1 (supporting info)

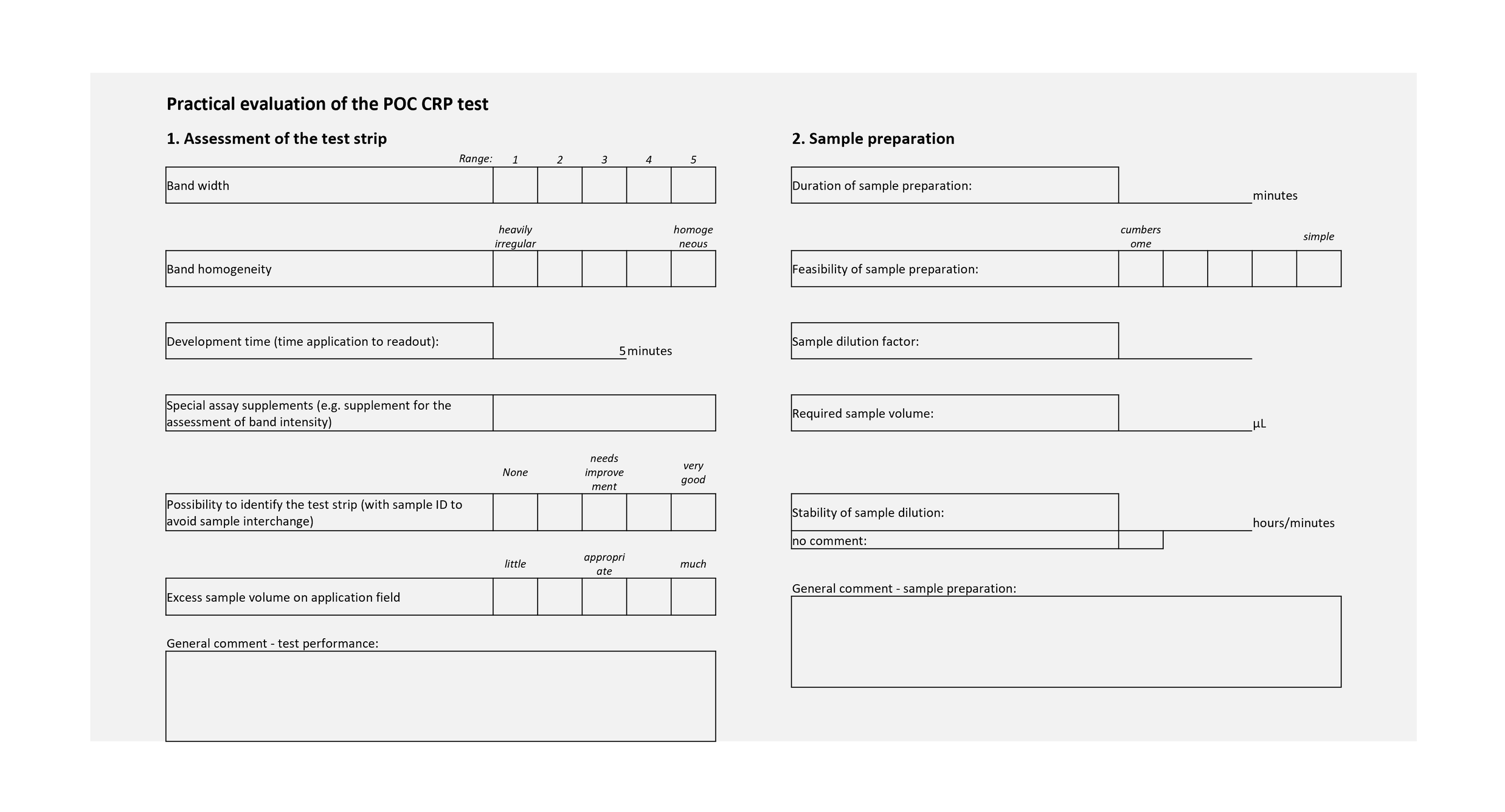
